# Dissociative experiences in fibromyalgia are mediated by symptoms of autonomic dysfunction

**DOI:** 10.1101/19006320

**Authors:** Daron Aslanyan, Valeria Iodice, Kevin A Davies, Hugo D Critchley, Jessica A Eccles

**Author notes:** CORRESPONDING AUTHOR: Dr Jessica Eccles, Trafford Centre for Medical Research, University of Sussex BN1 9RY.

## Abstract

**Background:** Fibromyalgia is characterised by chronic widespread pain. Quality of life is further reduced by autonomic and cognitive symptoms, including subjective ‘brain-fog’ and dissociative experiences. Although an association with joint hypermobility suggests variant connective tissue is a factor in both fibromyalgia and dysautonomia, the mechanisms underlying the neuropsychiatric symptoms are poorly understood.

**Methods:** 21 fibromyalgia patients and 21 healthy controls were assessed for joint hypermobility dissociative experiences, autonomic symptoms and interoceptive sensibility. Mediation analyses were conducted according to the method of Baron and Kenny.

**Results:** Patients with fibromyalgia reported greater dissociative experiences and autonomic symptoms. The relationship between fibromyalgia and dissociative experiences was fully mediated by symptoms of orthostatic intolerance. Fibromyalgia, dissociative experiences and orthostatic intolerance all were associated with joint hypermobility and interoceptive sensibility.

**Conclusions:** This exploratory investigation highlights the relationship between dissociative experiences in the context of fibromyalgia and subjective experience of aberrant physiological responses. These findings can enhance the recognition and management of neuropsychiatric symptoms in patients with fibromyalgia, wherein dissociative experiences reflect disturbance of self-representation that can arise through abnormalities in internal agency, autonomic (dys)control and interoceptive prediction errors.

## INTRODUCTION

Fibromyalgia is characterised by chronic widespread musculoskeletal pain lasting for more than 3 months ^1^ and has a prevalence in the UK population of approximately 2-5% ^2^. Patients with fibromyalgia score significantly lower in health-related quality of life domains than either those in the general population or patients with other musculoskeletal disorders such as rheumatoid arthritis ^3^.

Symptom domains that are associated with lower quality of life alongside pain include neuropsychiatric symptoms such as emotional fatigue, subjective and objective cognitive dysfunction (i.e. brain-fog) and dissociative experiences ^4^. These have recognised phenomenological overlap. Furthermore, the co-occurrence of brain-fog and dissociative experiences is associated with higher pain intensity and decreased mental well-being ^4^. Although patients frequently report these extra-musculoskeletal manifestations of fibromyalgia, the mechanisms underlying neuropsychiatric symptoms are poorly understood.

Many patients with fibromyalgia also experience symptoms of autonomic dysfunction (dysautonomia), particularly in association with orthostatic intolerance, which may reach criteria for Postural Tachycardia Syndrome (PoTS) ^5^. Fibromyalgia and dysautonomia are additionally both associated with connective tissue conditions specifically joint hypermobility ^6 7^. Joint hypermobility can be classified into generalised joint hypermobility (GJH) or and symptomatic joint hypermobility (Hypermobile Ehlers-Danlos syndrome, formerly known as Joint Hypermobility Syndrome/Ehlers-Danlos syndrome hypermobility type) in which GJH is accompanied with significant musculoskeletal or connective tissue symptoms. The Brighton Criteria are currently the only validated research criteria for diagnosis of symptomatic joint hypermobility ^8^.

This study aimed to investigate a possible mechanism for the basis of dissociative experiences in fibromyalgia, based on emerging theory linking the consciousness experience of integrity of self-integrity to the predictive control of internal bodily state ^9^.

## METHODS

### Participants

We recruited 22 patients with fibromyalgia from a Rheumatology Clinic in Sussex. Fibromyalgia was defined using modified 2010 The American College of Rheumatology (ACR) criteria ^1^. These participants were compared to 22 heathy controls recruited via advert free of major psychiatric or neurological disorder.

### Data collection

After collection of demographic and medical information, each participant underwent formal clinical assessment of joint hypermobility by a trained medical professional. Each participant then completed questionnaires examining dissociative and autonomic symptoms. In addition they completed self-report measure of interoceptive sensibility.

The Dissociative Experiences Scale-II (DES-II) ^10^ is the most commonly used tool to measure dissociative experiences and consists of 28 experience items respondents mark on a 11-point Likert scale.

An autonomic and quality of life self-administered questionnaire (AQQoL) ^11^ was used to quantify self-reported symptoms indicative of autonomic dysfunction across different domains, including orthostatic, gastrointestinal, bladder, secretomotor, sudomotor and sleep. Here we focus on orthostatic intolerance symptom severity (rather than impact on quality of life) as variable of interest because of the established link between fibromyalgia and orthostatic intolerance..

Interoceptive sensibility ^12^ was measured using the awareness subsection of the Porges Body Perception Questionnaire^13^.

Participants were assessed for presence of symptomatic joint hypermobility using the Brighton Criteria which incorporate the Beighton Scale ^8^.

Informed consent was received from all participants. The study was approved by the NRES Ethics Committee (South East Coast) (12.LO.1942)

### Statistical analysis

Anonymised datasets were analysed in SPSS (Version 25, SPSS Inc.).

Between-group differences in dissociative experience scores and symptoms of autonomic dysfunction were assessed using the Mann-Whitney U test as they were normally distributed. Between group differences in interoceptive sensibility scores were assessed using independent samples T test as these data were normally distributed. A statistical significance threshold of p <0.05 was used.

Analysis of whether orthostatic intolerance mediated the relationship between fibromyalgia and dissociative experiences was conducted according to the method of Baron and Kenny ^14^ and was adjusted for gender. Correlation analyses were conducted using linear regression.

This method necessitates that an independent predictor variable (fibromyalgia) is significantly and independently related to the mediator variable (orthostatic intolerance) and dependent variable (dissociative experiences). Full mediation is suggested if the relationship between the independent variable and dependent variable is rendered non-significant by addition of a mediator.

## RESULTS

### Characteristics of study population

After exclusion of 1 healthy control and 1 participant with fibromyalgia due to incomplete data, the study population included 21 patients with fibromyalgia (20 female; mean age 41.86 years) and 21 heathy controls (15 female; mean age 43.48 years). There were no significant differences in age between the two groups.

### Dissociative experiences

Patients with fibromyalgia reported significantly greater (mean, SEM) dissociative symptoms (48.67, 10.00) compared to controls (18.14, 2.72), (U= 134.0, p= 0.029).

### Autonomic dysfunction symptoms

Total autonomic symptom severity across all domains (mean, SEM) was higher in patients with fibromyalgia (83.48, 9.62) than controls (19.29, 2.49), (U=23.0, p=<0.001) and was significantly higher in each individual autonomic domain. Symptom severity in all atuonomic domains except bladder was significantly correlated with increase in dissociative symptoms. Orthostatic intolerance symptom severity explained 94% of the variance of total autonomic symptom severity.

### Symptoms of orthostatic intolerance

Patients with fibromyalgia reported significantly greater (mean, SEM) symptoms of orthostatic intolerance (41.90, 4.93) compared to controls (8.48, 1.06) (U=14.5, p<0.001)

### Analysis of relationship between fibromyalgia, dissociative experiences and orthostatic intolerance

It was shown that there was a significant correlation between fibromyalgia (independent variable) and dissociative experiences (dependent variable), which remained after adjusting for gender (β =0.406, p=0.011).

Correlation analysis, adjusting for gender suggested there was a significant relationship between fibromyalgia and orthostatic intolerance (β=0.693, p=<0.001) and between orthostatic intolerance and dissociative experiences (β=0.412, p=0.010).

Additionally, adjusting for symptoms of orthostatic intolerance rendered this relationship non-significant (β=0.231, p=0.281), suggesting full mediation (Figure 1).

**Figure 1:**
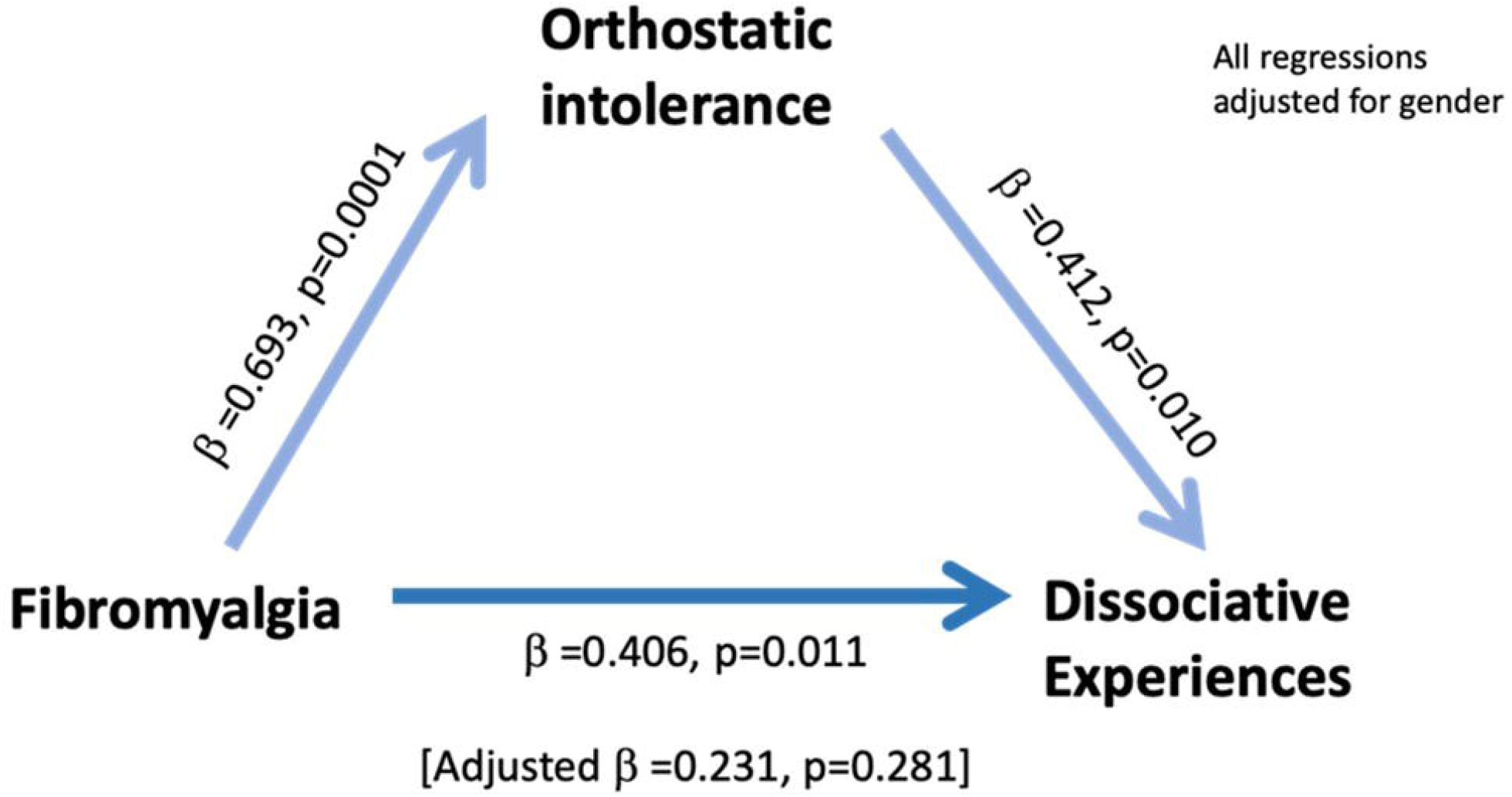
Graphic demonstrating full mediation of the relationship between fibromyalgia and dissociative experiences by symptoms of orthostatic intolerance

### Correlation with symptomatic joint hypermobility

All three variables (Fibromyalgia, dissociative experiences and orthostatic intolerance) correlated significantly with the presence of symptomatic joint hypermobility as defined by Brighton Criteria. Only 3 participants with Fibromyalgia did not meet Brighton Criteria, whereas only 6 controls met Brighton Criteria – this was significant (X^2^ 14.00, 1, p=<0.001). Patients meeting Brighton Critereria reported significantly greater (mean, SEM) dissociative symptoms (45.00, 3.15) compared to controls (17.94, 3.15), (U= 137.0, p=0.045). Patients meeting Brighton Criteria reported significantly greater (mean, SEM) orthostatic intolerance symptoms (36.04, 5.17) compared to controls (10.72, 1.82), (U=77.5, p<0.001).

### Relationship to interoceptive sensibility

Patients with Fibromyalgia reported significantly higher (mean, SEM) interoceptive sensibility (121.15, 5.76) compared to controls (94.55, 9.90), (t=2.32, df 38, p=0.026).

## DISCUSSION

This is the first study, to the authors’ knowledge, that explores dissociative experiences in the context of fibromyalgia in relation to autonomic symptoms. The results are significant suggesting, for a patient population vulnerable to neuropsychiatric symptoms, dissociative experiences are mediated by symptoms of orthostatic intolerance.

The results reinforce existing evidence for the known associations between fibromyalgia and dysautonomia ^5^ and between fibromyalgia and dissociative experiences ^4^. Importantly, this is the first account to explicitly link all three phenomena.

Previous work has suggested that hypermobility, a common connective tissue condition, has a multisystem phenotype and its presence is a vulnerability factor for a variety of neuropsychiatric symptoms ^15^. Our study suggests that abnormalities of connective tissue found in joint hypermobility disorders and linked to fibromyalgia may also fundamentally underlie the expression of autonomic dysfunction and, in turn, predispose to dissociative experiences.

These results offer fresh avenues to enhance the formal recognition and treatment of cognitive symptoms including dissociative experiences and brain-fog, which can be debilitating and significantly lower quality of life ^4^. Therapeutic methods to alter both autonomic function itself and how autonomic changes are subjectively experienced may be useful approaches to the management of these cognitive symptoms. Nevertheless, this small study indicates a need for more research to develop and fully characterise these results. In addition to larger sample sizes, further studies would usefully combine objective autonomic testing, a more representative gender mix, updated definitions of hypermobility syndromes, and a broader set of measures to differentiate dissociative experiences.

Significantly, our results are also in line with theoretical advances in consciousness science that hypothesise a basis to dissociative experiences in abnormalities of self representation arising through aberrant internal agency ^9^. This model is based on Bayesian neural computations and the signalling of interoceptive prediction error, which may be revealed by subjective symptoms of orthostatic intolerance. Of note patient participants had enhanced interoceptive sensibility.

In conclusion, this exploratory study of fibromyalgia points to a mechanism behind commonly reported neuropsychiatric symptoms; our results indicate that subjective symptoms of orthostatic intolerance mediates the relationship between fibromyalgia and dissociative experiences.

## Data Availability

Deindentified participant data is available upon reasonable request from the corresponding author (ORCID ID https://orcid.org/0000-0002-0062-1216).

## Acknowledgments

We would like to thank Lorraine Shah-Goodwin and Zdenka Cipinova at the Clinical Investigation Research Unit, Brighton and Sussex University Hospitals NHS Trust for their assistance in data collection and recruitment.

## Data availability

Deindentified participant data is available upon reasonable request from the corresponding author.

## Contributorship statement

JAE, HDC, KAD and VI designed the work. DA and JE analysed and interpreted data and drafted the article. All authors revised the work and agreed on the final version. Data was collected by LS-G and ZC.

## Patient Consent

All participants gave informed consent to take part in the research

## References

1. Wolfe F, Clauw DJ, Fitzcharles MA, et al. Fibromyalgia criteria and severity scales for clinical and epidemiological studies: a modification of the ACR Preliminary Diagnostic Criteria for Fibromyalgia. J Rheumatol 2011;38(6):1113–22. doi: 10.3899/jrheum.100594 [published Online First: 2011/02/01]

2. Jones GT, Atzeni F, Beasley M, et al. The prevalence of fibromyalgia in the general population: a comparison of the American College of Rheumatology 1990, 2010, and modified 2010 classification criteria. Arthritis Rheumatol 2015;67(2):568–75. doi: 10.1002/art.38905

3. Verbunt JA, Pernot DH, Smeets RJ. Disability and quality of life in patients with fibromyalgia. Health Qual Life Outcomes 2008;6:8. doi: 10.1186/1477-7525-6-8 [published Online First: 2008/01/22]

4. Leavitt F, Katz RS, Mills M, et al. Cognitive and dissociative manifestations in fibromyalgia. J Clin Rheumatol 2002;8(2):77–84.

5. Staud R. Autonomic dysfunction in fibromyalgia syndrome: postural orthostatic tachycardia. Curr Rheumatol Rep 2008;10(6):463–6.

6. Sendur OF, Gurer G, Bozbas GT. The frequency of hypermobility and its relationship with clinical findings of fibromyalgia patients. Clin Rheumatol 2007;26(4):485–7. doi: 10.1007/s10067-006-0304-4 [published Online First: 2006/04/25]

7. De Wandele I, Calders P, Peersman W, et al. Autonomic symptom burden in the hypermobility type of Ehlers-Danlos syndrome: a comparative study with two other EDS types, fibromyalgia, and healthy controls. Semin Arthritis Rheum 2014;44(3):353–61. doi: 10.1016/j.semarthrit.2014.05.013 [published Online First: 2014/05/14]

8. Grahame R, Bird HA, Child A. The revised (Brighton 1998) criteria for the diagnosis of benign joint hypermobility syndrome (BJHS). J Rheumatol 2000;27(7):1777–9.

9. Seth AK, Suzuki K, Critchley HD. An interoceptive predictive coding model of conscious presence. Front Psychol 2011;2:395. doi: 10.3389/fpsyg.2011.00395 [published Online First: 2012/02/01]

10. Bernstein EM, Putnam FW. Development, reliability, and validity of a dissociation scale. J Nerv Ment Dis 1986;174(12):727–35.

11. Iodice V. The Postural Tachycardia Syndrome: a multi-system condition. Clinical features, pathophysiology, genetics and novel treatment, 2013.

12. Garfinkel SN, Seth AK, Barrett AB, et al. Knowing your own heart: distinguishing interoceptive accuracy from interoceptive awareness. Biol Psychol 2015;104:65–74. doi: 10.1016/j.biopsycho.2014.11.004 [published Online First: 2014/12/03]

13. Porges S. Body perception questionnaire. Laboratory of Developmental Assessment, University of Maryland 1993

14. Baron RM, Kenny DA. The moderator-mediator variable distinction in social psychological research: conceptual, strategic, and statistical considerations. J Pers Soc Psychol 1986;51(6):1173–82.

15. Eccles JA, Beacher FD, Gray MA, et al. Brain structure and joint hypermobility: relevance to the expression of psychiatric symptoms. Br J Psychiatry 2012;200(6):508–9. doi: 10.1192/bjp.bp.111.092460 [published Online First: 2012/04/26]

